# Auricular Muscle- controlled Navigation for Powered Wheelchairs

**DOI:** 10.64898/2026.02.28.26347311

**Authors:** Aleksandra Nowak, James Fleming, Massimiliano Zecca

**Affiliations:** Wolfson School of Mechanical, Electrical and Manufacturing Engineering, Loughborough University, Loughborough, United Kingdom

## Abstract

There are many alternative methods to joystick control for control of Electric Powered Wheelchairs for users with neuromuscular disabilities, such as muscular dystrophy, and spinal cord injuries, such as tetraplegia. However, these methods- which include the sip-and-puff method, head and neck movement, blinking, or tongue movement- hinder social interaction, and are therefore detrimental to user independence. In recent years, research has explored the use of Electromyography (EMG) signals from alternative muscles to control a powered wheelchair, consequently increasing the quality of life of these users. The Auricular Muscles (AM) may be suitable, as they are controlled separately from the facial nerve and are vestigial in humans, making them advantageous for powered wheelchair control for users with tetraplegia. Additionally, they are located around the ear, adding a level of cosmesis when designing wearable sensors and prosthesis. This paper extracts and implements two control strategies from current literature and, for the first time, compares them directly, demonstrating viable implementation approaches for an online EMG-based powered-wheelchair control system. A Support Vector Machine (SVM) was developed and various window lengths were compared, with the most accuracy and real-time effectiveness found at 300ms. A study with three participants demonstrates the feasibility of these methods of control as well as experimental results to guide the potential AM use.

## Introduction

Currently, many Human–Machine Interfaces (HMIs) exist as alternatives to conventional joysticks or single-switch controls for powered wheelchairs, aimed at users with motor disabilities, such as those resulting from spinal cord injuries [1]. For these users, it may not be possible to control powered wheelchairs using conventional control due to level of injury, pain, or impairment. Current alternative methods include the sip-and-puff method [1], blinking [2], neck movement [3], tongue movement [4] and blink and bite [5]. However, despite their controllability and functionality, they come with limitations. In a survey by Fehr et al. [6], clinicians approximated that 40% of users find it difficult or impossible to use their wheelchair for steering and manoeuvring tasks, signifying that current technology in the area is inadequate. Also, they hinder social interactions [7], and do not adequately meet the needs of specific user groups, such as individuals with various spinal cord injuries and muscular disorders [6].

Furthermore, users with spinal cord injuries report experiencing depression and a diminished quality of life [8], highlighting the limitations of current assistive technologies in fully supporting their independence and well-being. Various research has been conducted into alternative methods of control of a powered wheelchair for users with limited ability to control or input instructions [9], which include users with the neuromuscular disease, Amyotrophic Lateral Sclerosis, as well as the progressive genetic muscular disorder, Muscular Dystrophy, and victims of stroke or spinal cord injuries.

An explorative method of control for these users in recent years has been using EMG sensors on more accessible muscles that are less affected by the disability of the user [10, 11]. EMG signals are electrical signals passing through muscles and can be measured during their contraction and relaxation for control of a prosthetic limb or powered wheelchair [12]. This is an appealing method of control, as it could be tailored to individuals depending on their condition, disability, or the current state in progression of an illness.

The Auricular Muscles (AM) have been identified as a group of muscles of interest, as their use would provide a method of control that interferes with social interactions the least, since the AM are vestigial muscles, no longer used by humans, and their original function may have evolved [13, 14]. Utilizing these muscles would allow users to control assistive devices [15–17] while performing other activities, such as talking, or neck movement. With their concealed nature, any wearable devices would not as intrusive as current control methods, increasing the cosmesis and potential confidence of the wearer.

The AM would be particularly promising for users with tetraplegia or similar conditions, as they are not controlled by the facial nerve [18], meaning individuals with disabilities who retain facial movement abilities could operate an AM-based system without inhibiting their daily lives and social interactions, therefore enhancing their overall quality of life. The AM have been investigated for several other applications. For example, for the monitoring and alleviation of Post-Traumatic Stress Disorder symptoms [18], cursor control [17, 19], control of a robotic thumb [20], prosthetic arm [21] and auricular acupuncture for immediate relief from acute lumbar sprain pain was investigated by Tang et al. [22]. Additionally, the AM have been researched for use of control of assistive devices, such as powered wheelchairs [13]. There are recent studies which investigate control of powered wheelchairs in response to the EMG signals generated from the auricular muscles [16].

This work implements two control strategies from the literature-namely the Continuous Control Strategy [16] and the Morse Code Wheelchair Navigation [23] strategies-to navigate around a small maze wirelessly using a small robotic prototype. These two strategies were found to be most safe and intuitive, in a previous study by the authors [24]. Three participants were recruited for the study, which provides evidence that the two strategies may differ in suitability for specific user groups— such as tetraplegic or post-stroke users—and presents a novel implementation method designed for fast, real-time signal classification.

A classifier was developed for the two strategies, and window sizes were compared with the most optimal time window found to be around 300ms. However, we found that depending on the control strategy on the AM, this may vary. This kind of practical comparison and adaptation of window lengths for an EMG-controlled assistive device, has not been done before.

## Related Work

A large amount of research effort has been made in developing new HMI for powered wheelchairs, as well as their comparisons; indicated by the survey by Ghorbel et al. [1], which overviews methods such as neck movement, sip-and-puff, and blinking. Research into assistive technology is growing, and particular interest is in EMG [25]. Fehr et al. [6] approximated 40% of users would benefit from the development of new controller interfaces to accommodate their needs and abilities. Since then, more and more research has been done in the area, especially in EMG control [25].

Few studies have explored the use of the Auricular Muscles (AM) for wheelchair control, representing a clear gap in the literature. Investigating AM-based control is important, as it could provide an alternative method for steering powered wheelchairs, enhance independence, and improve quality of life for users with tetraplegia, addressing challenges highlighted by clinicians in Fehr et al. [6].

### EMG Signal

EMG signals can be measured intrinsically (imaging-EMG) using fine needle insertion, and extrinsically (surface-EMG) using surface electrodes [25]. While imaging-EMG provides detailed information on muscle fibre activity, a broad view of the muscle transient state can be adequately captured using surface-EMG [25], so for the purpose of the study, surface-EMG acquisition will be used.

In general, the EMG signal suffers from several problems. Firstly, it is small in amplitude-the raw surface-EMG signal is normally found to be between 0 to 10mV peak-to-peak [26] hence, it requires amplification. Secondly, it suffers with noise pollution and interference, as electrical noise (50-60Hz) is picked up [26]. Thirdly, motion artefact-involuntary movement of the equipment or wire-introduces low-frequency noise, in the range of 0-20Hz [26]. Lastly, repetitive and continuous use of muscles (muscle fatigue) will lead to changes in surface-EMG amplitude. For example, Xu et al. [27] measured that the amplitude of surface-EMG signals without fatigue is under 400uV and rises to 600uV as muscle fatigue occurs, as well as the mean and median spectral frequency decreasing 10.8% and 17.2% respectively after 60mins of muscle contraction. Due to these problems, appropriate features and classification techniques should be considered when designing a myoelectric-based system [28].

### EMG Classification

EMG signals may be identified using the threshold method, or using classifiers. However, Xu et al. [27] state that surface-EMG patterns will change as muscle fatigue occurs, meaning the threshold method will become less effective.

To counter these issues, signal processing techniques such as filtering and amplification are applied to isolate the signal. Extraction of features such as the amplitude, duration, and RMS from the time domain, and frequency characteristics of the signal from the frequency domain takes place to correctly identify the EMG signal from noise and motion artefact [29].

Due to the nature of the signals, amplifiers used may vary between a gain of 1000 for the forearm muscles [30] to 5000 for muscles on or around the shoulder [25], or sometimes high resolution data is simply passed through an ADC instead of using a high-gain amplifier [25].

The usable range of EMG signals is between 0 and 500Hz, so according to the Nyquist-Shannon sampling theorem [25], which states that the sampling frequency should not be less than twice the bandwidth, a sampling frequency of around 1000-1024Hz is often used. However, some studies use lower rates of 125, 128, or 512Hz depending on the application and implementation [25].

Furthermore, Cimolato et al. [31] conducted a systematic review of EMG controlled lower limb prosthesis, and from this survey of 56 papers, within the digital signal processing of the systems, 36 papers included bandpass filters from 10-30Hz to 350-500Hz (with one paper ranging from 20-1000Hz), and another 6 included a High Pass Filter (HPF) and a Low Pass Filter (LPF) in similar ranges. The remaining papers included either one LPF or HPF or no filter, or filter information was not specified. Often, notch filters around 50 or 60Hz are also used to remove the ambient electrical noise of power-line interference [25]. However, since the dominant energy of the EMG is located within the 50-100Hz range, filtering at these frequencies can attenuate important signal components and is therefore advised against in some instances [26].

To classify the signal, in the pattern-based method of control, the Support Vector Machine classifier is one of the most common and most recommended by literature [25]. Other successful examples include Fuzzy Logic-based classifiers and Artificial Neural Networks. Non-pattern recognition method used include Finite State Machines [25].

To control a powered wheelchair with EMG, Iqbal et al. [32] compared several classifiers and achieved an accuracy of 99.05% when classifying hand gestures using 8 channels with multi-channel SVM. In trials with six able-bodied participants, all were able to control the wheelchair without collisions, demonstrating real-world usability alongside high accuracy.

Phinyomark et al. [33] claim that EMG features based on frequency domain are not good in EMG classification. Hudgins’ set of features is widely used and proven in several studies [34].

Englehart and Parker [35] proposed an alternative wavelet-based classification approach to Hudgins’ time-domain one. Using data from 16 healthy subjects and flexion/extension of the elbow and pronation/supination of the forearm, they yielded 0.5% error when discriminating four classes, significantly lower than Hudgins’ 10%.

### Control Methods

Additionally, there are many various methods of control for a powered wheelchair, as a sublayer of the HMI. However, it has been found that the minimum practically usable number of control commands is five: forward, backward, turn left, turn right, and stop [25]. So, all EMG-based control strategies should cover these instructions.

Huda et al. [36] proposed a unique mathematical approach for real-time hand gesture recognition to control a smart wheelchair using only finger movement. They used an RGB camera to detect three gestures: ‘Drive’, ‘Stop’ and ‘Horn’, and when in driving mode, a finger is tracked and wheelchair movement is performed as per the direction of that finger.

A study by Zhang et al. [5] differentiated voluntary blink and bite patterns at an accuracy of 92.6% to control a powered wheelchair. Zhang et al. [2] expanded on this work and tested a system based on morse code blinking for both control of home appliances and communication. The system measured surface-EMG on the corrugator supercilii muscles, integrated into a wearable glass set, which claimed to improve the training experience.

Xu et al. [27] used single and double jaw clicks to navigate a wheelchair around a 6×8m course with 2 obstacles. The proposed HMI system performed well, despite the occurrence of muscle fatigue increasing the time to complete the course from 103s to 154s. The study suggested future work should remove noise signals from facial movements when a user is talking or moving around, as these would interfere with social interactions and use of the system.

Furthermore, Schmalfuß et al. [16], implanted their own self-constructed minimally invasive electrodes in the Posterior AM in two individuals with spinal cord injury to control a powered wheelchair. The two subjects were one ear-wiggler and one non-ear wiggler. Though the dropout rate of the intensive study was high, quantitative results were promising-with mean reaction times decreasing from 585ms to 408ms in contraction and 692ms to 462ms in relaxation suggesting that subjects learnt to control their Posterior AMs faster throughout the study. Correct activation also doubled from 26.7% to 50.6%, proving that control of the Posterior AMs can be improved with training. However, the subjects estimated that they would need 7 to 14 days more of training to achieve safe and complete control of the wheelchair using this method, demonstrating the potential for further work.

Mentioning this work, Pinheiro et al. [15] trained subjects to carry out a 2D task on a monitor using their Superior AMs. Subjects achieved a final success rate of 72.22%, an improvement from the initial 52.78% median value. The study implemented the use of thresholds, with a contraction below 35% of the Maximum Voluntary Contraction (MVC) of the right AM moving the cursor left, and a contraction of 35-70% MVC moved the cursor right (similar logic for up and down on the left AM). Over 70% was used to toggle the locked movement state but would have to be activated for 500ms to activate this state. Additionally, a neural network was trained on the subjects’ facial expressions, and a penalty was added in the tasks to discourage users from using facial expressions to control their AM. However, the study did not compare any other approaches to control the cursor using the AM to assess the most intuitive one.

Manero et al. [11], suggested the ‘Uni-Lateral’ and ‘Bi-Lateral’ methods of wheelchair control using the temporalis muscles on the forehead, named after the activation of singular or both muscles respectively, for users with Amyotrophic lateral sclerosis. The method uses the differentiation of long and short surface-EMG signals from the muscles, as well as strong and small signals, to navigate using a wheelchair. Four participants with slowly progressive Amyotrophic lateral sclerosis trialed the system, with reverse control initially being disabled for 2/4 of the participants on the first trial and later disabled for all participants. Unfortunately, the study did not collect timing data.

The Continuous Control strategy (CCS) implemented by Schmalfuß et al. [16] worked well, with the study resulting in all participants able to steer a wheelchair with EMG on both AM sides. A constant contraction on the left side corresponded to activation of the right motor- and a left turn. As a constant contraction on the right will turn right. Simultaneous contraction of both sides at the same time will drive the wheelchair forward. No contraction-no input-will mean no movement, and a stop of the wheelchair.

Furthermore, the study suggested the implementation of reverse driving capability through a double-quick contraction. While Schmalfuß et al. [16] did not test this themselves, it has been implemented within this study to increase the capability of this control system strategy, as well as to satisfy the requirement that all powered wheelchairs should have 5 instructions, which includes the reverse [25]. In CCS, a double contraction will switch the robot to ‘reverse mode’ from which the usual control applies, but in reverse-left and right contractions move the robot in reverse left and right directions respectively.

The Morse Code Wheelchair Navigation (MCWN) implemented by Ka et al. [23] offers an alternative method of control, possibly where the muscles for control are limited, as it requires just one muscle which can be flexed for long or short durations. The strategy was designed on a switch: a long signal followed by short cause a 90-degree left turn. Two long pulses cause a 90-degree right turn. Two short pulses drive the wheelchair forward. A short signal followed by a long will cause reverse driving. This has been translated to be used with EMG, and has been implemented in this study, as it would offer an interesting comparison to the other strategy, which was specifically designed for control with the AM.

In previous work [24], the Uni-Lateral and Bi-Lateral strategies implemented by Manero et al. [11] were also both compared using the Flexor carpi radialis muscles, but results showed that participants favoured the MWCN and CCS the most, therefore these have been now implemented on the AM and compared in this study.

## Materials and methods

The MyoWare 2.0 Muscle Sensor was chosen for its pre-implemented filters, consisting of an instrumentation amplifier of gain 200, and bandpass filter 20-498Hz. The sensor was used in Envelope mode which rectified the signal, and passes it through a LPF envelope detection circuit. The envelope *e*(*t*) may be given as:

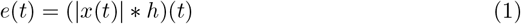

Where * is the convolution and *h* is the impulse response of a low pass filter. The frequency cut-off for the LPF is 3.6Hz, as it provided the highest resolution and features for the classifier. The signal was then sampled at 47Hz to satisfy the Nyquist theorem (over 7.2Hz) and provide enough resolution (with oversampling), before being passed through an SVM classifier. The control strategies were implemented on the output of this, in order to control the prototype. To avoid grounding and power supply problems, as well as lack of hardware to support multiple wired sensors, the MCWN was run on a wired sensor, and the CCS was run on two wireless sensors. Both underwent the same signal processing and had the same sampling frequency.

For the purpose of testing the various control strategies, the maze was re-constructed of size 1.8×3.0m using wood and aluminium brackets, as seen in Fig 1.

**Fig 1.**
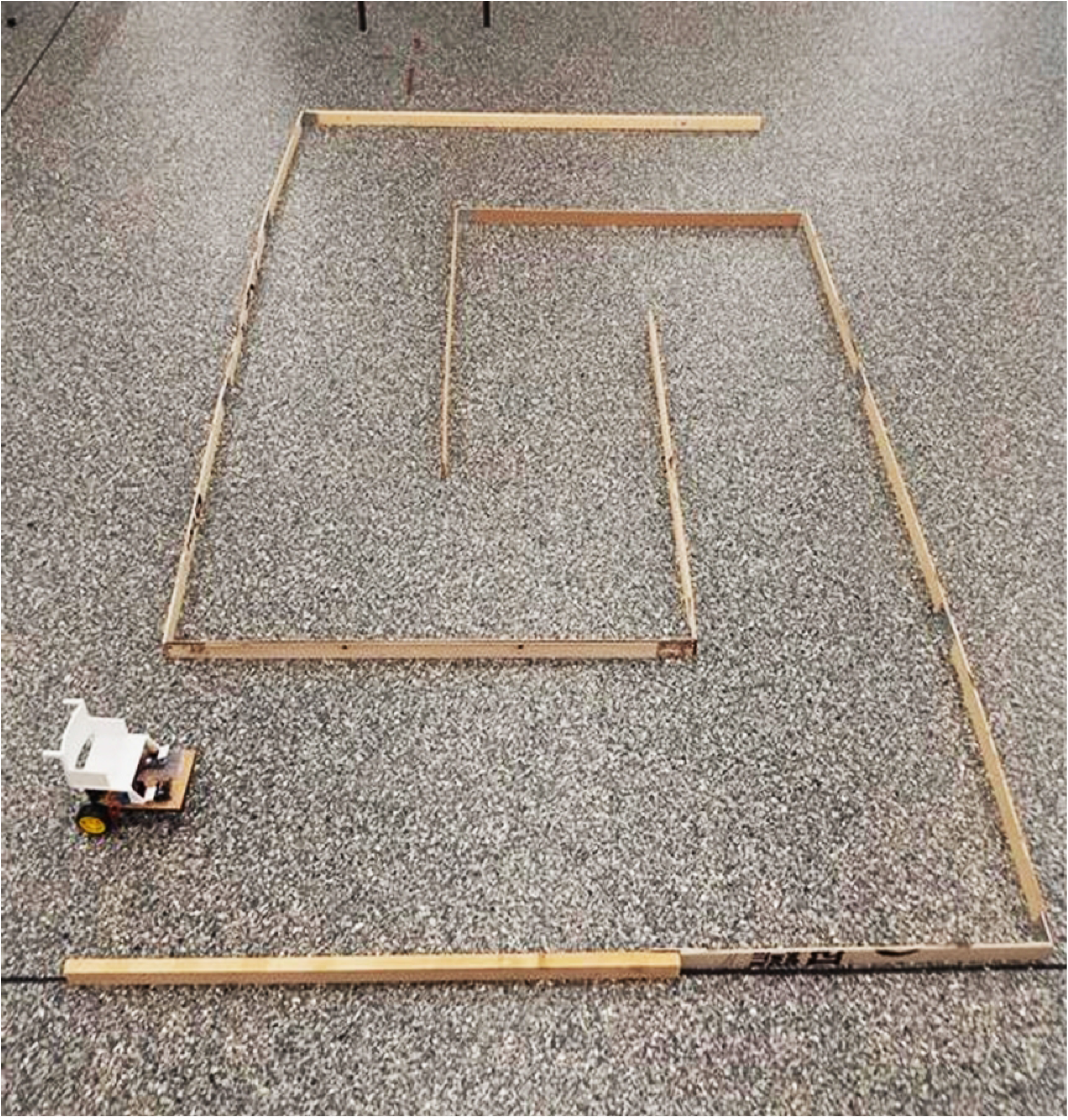
Maze and Prototype setup. 1.8×3.0m maze of equal width corridors secured to the floor with tape. Robotic prototype with local proportional control is controlled wirelessly using BLE protocol.

Many studies have been made comparing different window sizes to classifier accuracy. It has been generally agreed that a sliding window should be a maximum of 300ms, otherwise it would compromise usability [30]. Some studies even suggest that windows of 32ms [37] work well enough. In this study, several window sizes were compared and 300ms was chosen to be the most optimal, as it was considered the best trade-off between high classifier accuracy and a short time window.

The dataset for the construction of the classifier was divided into three segments: the Training set, Validation set, and Testing set, where all sets of data contained long signals, short signals and noise. A train/validation/test split of 80%/6.7%/13.3% was used, with the validation set used for hyperparameter tuning and the test set used to assess generalisation to unseen data. The accuracy on the testing dataset is the final classifier accuracy. The data was taken in 30s samples of mixes of long, short, and noise signals. Following manual data labelling in MATLAB, the data was segmented according to the selected window size, and then features extracted for each segment. The classifier was trained with Scikit-Learn 1.5.2 library in Python 3.10.2.

### Feature Extraction

Purely frequency domain features were not further explored in the classifier, as they do not provide a high enough accuracy to justify the computational cost, and are considered unnecessary, as literature shows that a high accuracy may be achieved using only time and wavelet features [33, 35]. Feature reduction took place by analysing the correlation between features. Overall, all thirteen features were used for the classification of signal/no signal. However, to reduce the time of processing, only Variance, RMS, Range, Kurtosis, Maximum and Average and Total slope changes, Skewness were used for fast classification under 100ms-used for MCWN second layer classification.

The following features are computed from the EMG signal over a fixed analysis window. Throughout this section, *x*_*i*_ denotes the *i*-th sample in the window, and *N* denotes the total number of samples in that window. The sample mean is denoted by 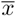, Where 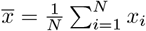, and the sample standard deviation is denoted by *σ*, where 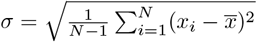.

### a) Total Sum

The sum of all signal values over the analysis window. It reflects the overall amplitude accumulation of the signal.

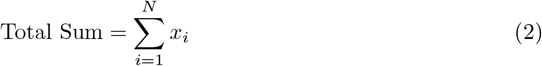

### b) Mean Absolute Value (MAV)

The average of the absolute values of the signal, providing a measure of overall signal magnitude regardless of direction.

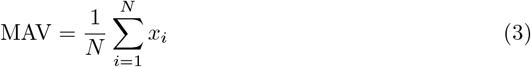

#### c) Square Integral (SI)

The sum of squared signal values, representing signal energy.

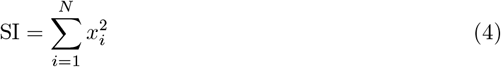

#### d) Variance

Measures the dispersion of the signal around its mean, indicating variability.

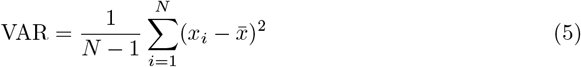

#### e) Root Mean Square (RMS)

The square root of the mean of squared values; reflects the power of the signal.

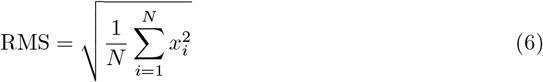

#### f) Range

The difference between the maximum and minimum values of the signal.

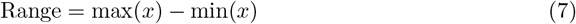

#### g) Kurtosis

A measure of the prominence of a distribution’s tails relative to its overall variance, useful for detecting transients or outliers.

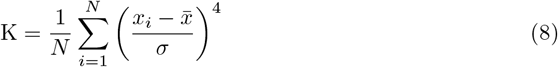

#### h) Maximum Slope Change

The largest difference between consecutive samples, representing the steepest signal transition.

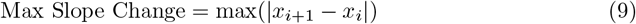

#### i) Average Slope Change

The mean of absolute differences between consecutive signal samples.

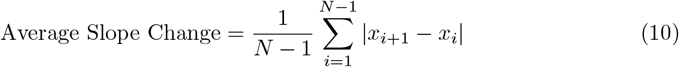

#### j) Total Slope Change

The sum of absolute differences between adjacent samples over the signal.

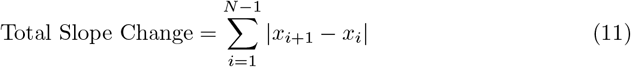

#### k) Skewness

Skewness measures the asymmetry of an EMG signal’s amplitude distribution, indicating whether the signal leans more to the left or right of the mean.

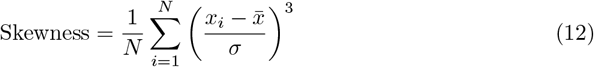

#### l) Wavelet Transform Threshold

A nonlinear feature derived from the denoising process using continuous wavelet transform using complex Morlet wavelet [38] given by:

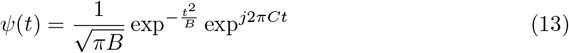

Where *B* is the Bandwidth parameter 1.5, and *C* is the Centre frequency 1. The signal is decomposed into wavelet coefficients, and coefficients below a threshold (often based on noise level) are suppressed. The remaining energy or count of coefficients above the threshold is used as a feature. For this feature, the coefficient at half time of the sample at 2Hz frequency is extracted.

### SVM

An SVM with a Radial Basis Function (RBF) kernel was used, with the hyperparameters C (Regularisation Parameter) and gamma (Kernel Coefficient) tuned manually to minimise misclassification on the validation set. C controls the trade-off between maximising the margin and minimising classification error. A high C value is less tolerant to misclassification. This means a high C value gives a higher accuracy but causes the model to generalise less. Gamma controls the shape of the decision boundary via the kernel. A low gamma risks underfitting, and a high gamma risks overfitting.

Our radial basis kernel function for determining the similarity of two vectors is given as follows:

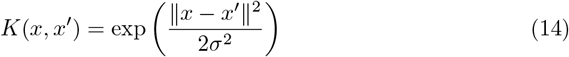

where 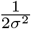 is our hyper parameter gamma.

We then use this function in our SVM classifier [39] which is defined as follows:

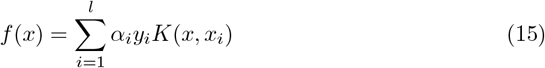

Where each *x*_*i*_ is a support vector learned from training, *y*_*i*_ that support vector’s label, *α*_*i*_ the weight associated with that vector, *l* the number of support vectors and *f* (*x*) the predicted label for input data *x*.

The SVM was created to identify the EMG signals, and Fig 2 shows the accuracy plotted against window size for the binary (signal/no signal) classifier. The ideal window size is therefore assumed to be 350-400ms.

**Fig 2.**
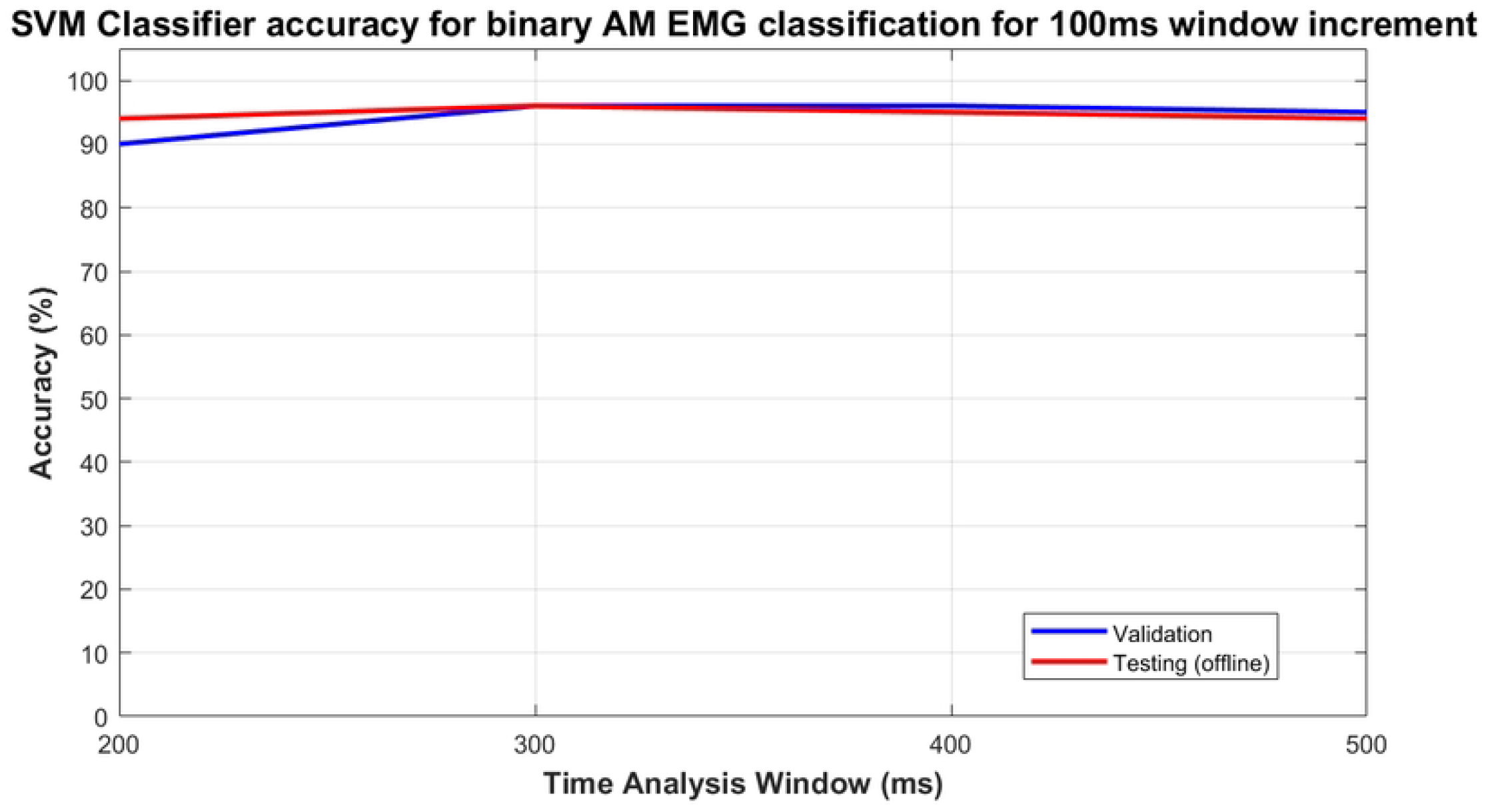
Classifier accuracy for signal. Classifier Validation and Testing Accuracies for binary signal detection classifier at 100ms time analysis window increments

A second classifier was developed to categorise long against short signals: removing everything else as noise. The accuracy against window size may be seen in Fig 3. It can be seen that accuracy improves with window length. This may be because a long signal is approximately 1s long, whereas a short signal is approximately 0.3s in duration.

**Fig 3.**
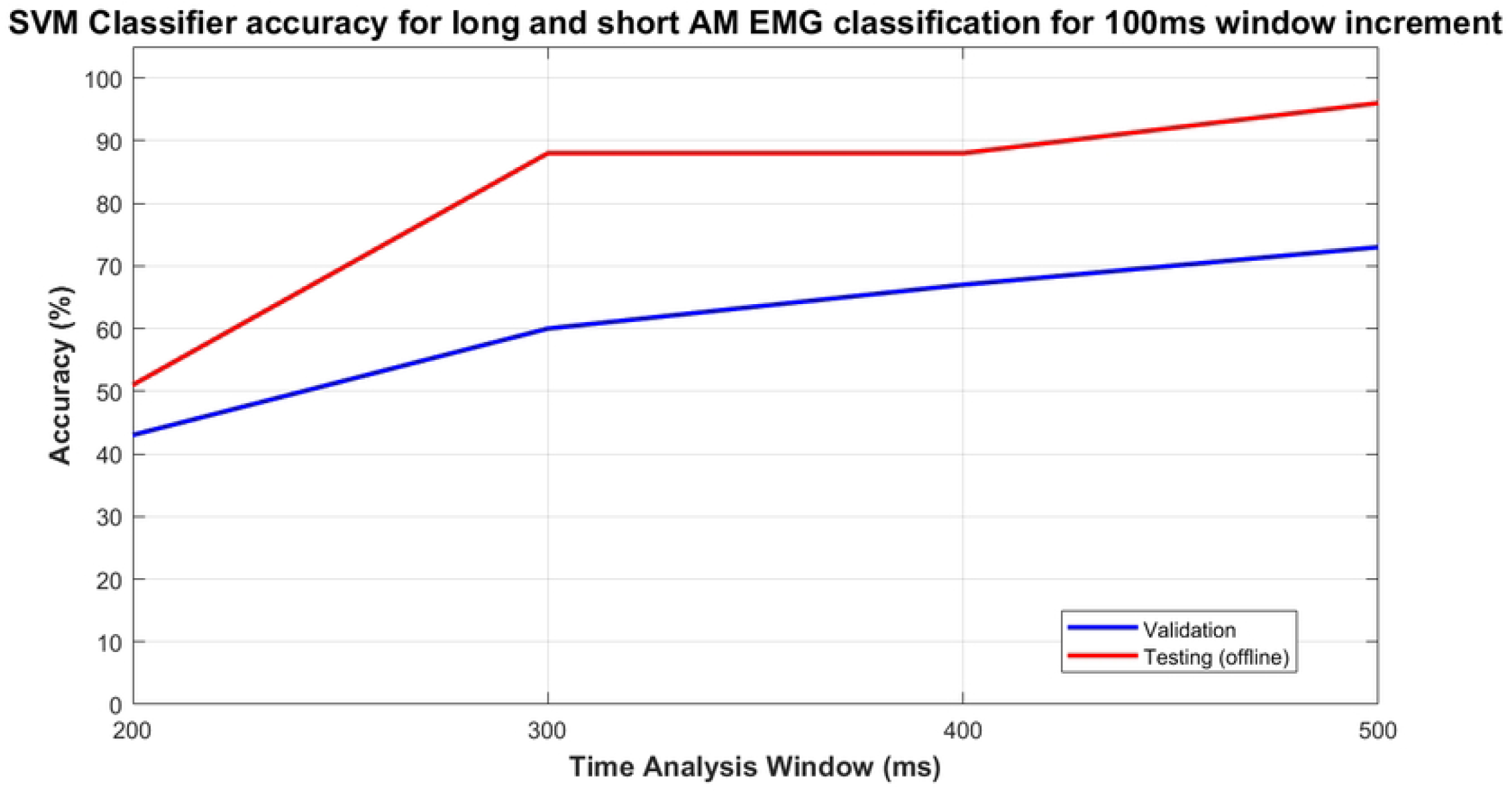
Classifier accuracy for long vs. short. Classifier Validation and Testing Accuracies for binary long/short classifier at 100ms time analysis window increments

### Control Strategies

#### 0.0.1 Continious (CCS)

For CCS, a classifier trained only on the first initial incline of the signal (first 200ms) is used to identify if a signal is coming in. Then, a thresholding method is used to determine if the muscle is still being activated. If a right muscle is being activated, without nothing being detected from the left side, the robot should turn right for as long as it is activated. If a left and right are detected at close enough times, the robot will move forward until one or both signals drop out permanently. If a left signal is detected, and it is a short length, the system will wait for a short time to see if another short left is sent (this activates the toggle reverse function), otherwise it will execute the short left turn with a delay, logic visible in Fig 4.

**Fig 4.**
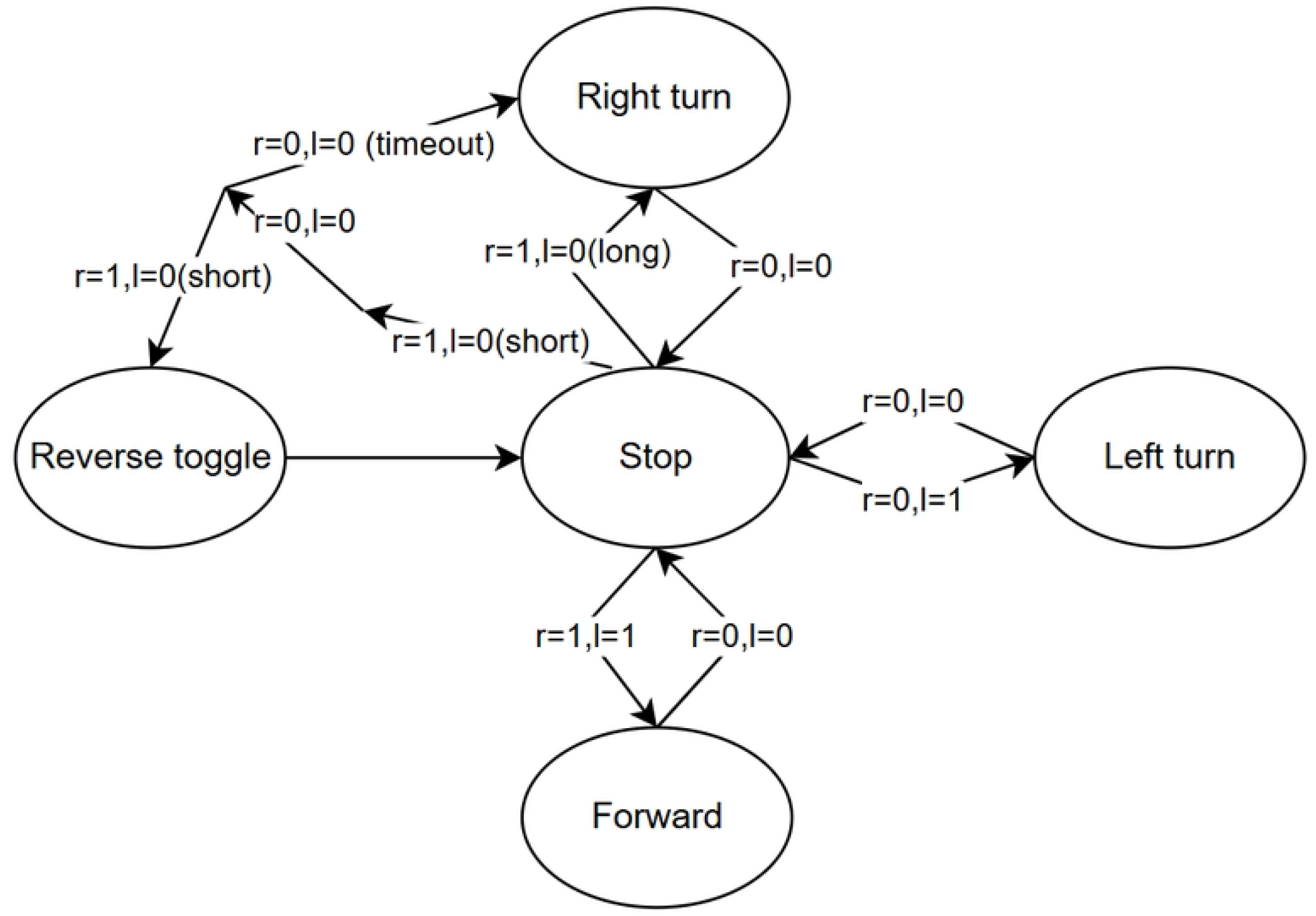
Finite State Machine for CCS. An ‘r=1’ means a high from the right, and ‘l=1’ mean a high from the left side. ‘0’ means no signal detected.

#### 0.0.2 AM Morse Code (AM-MCWN)

For AM-MCWN, it was found that the most effective classifier was a two level SVM, as it produced a higher real-time accuracy than a 3-class SVM (one-vs-one). The first level identifies the presence of a signal, and the second level identifies if it is long or short, logic visible in Fig 5.

**Fig 5.**
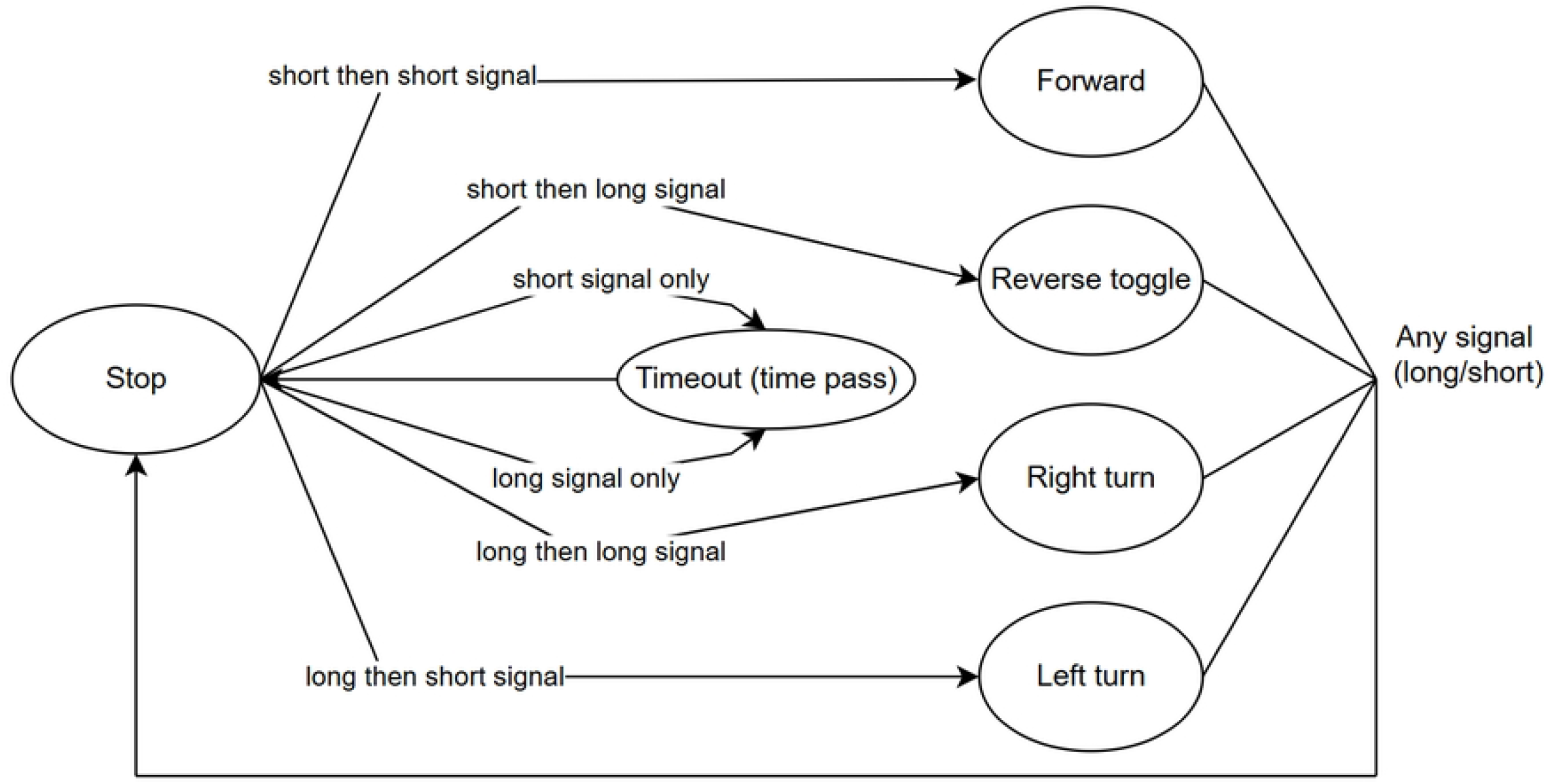
Finite State Machine for MCWN. Arrows show transitions between states.

Several improvements have been made to the original MCWN by the authors of this study to adapt the strategy to the AM and increase usability. Specifically, the forward/backwards, right back/forward turning and left back/forward turning functions can be interrupted by a stop function in the form of any signal: short or long.

Two buffers have been implemented for the AM-MCWN logic. The first buffer (300ms) is used for the signal/non signal classification. If a signal has been detected, a longer buffer (500ms) is used for identification of long or short. Both buffers are implemented as threads in Python; therefore, the first 300ms of the 500ms buffer is identical to the first, then fills up after a computation delay.

As a buffer is used to listen to both signals (each instruction is formed of two signals), if a function is being executed, the system does not wait for the stop to travel through the buffer, nor does it pass it through the second long/short classifier, it simply stops. This has reduced the stop of the robot from 2s to approximately 200ms. Additionally, for erroneous inputs, such as a short contraction when it was meant to be long, the system will timeout after 2s, and discard that input, increasing the useability and safety of the system.

## Results

The updated system, which utilises the Posterior Auricular Muscles, was tested on three healthy participants aged 18-24. This was conducted in accordance with the ethical approval of Loughborough University Ethics Committee (ethics ID: 20634).

All three participants could describe themselves as ear-wigglers, or able to move their ears. Only one of the participants (P3) was able to move their ears independently of each other.

The testing was done in two sessions, where the participant was plugged into the system, offered 5-10 minutes to practice using the system, before a run-through of the maze was completed with a control strategy. CCS and AM-MCWN were tested, one per session. Participant’s final run time, number of instructions the system detected and the number of crashes the prototype had with the walls were recorded, and can be seen in Table 1. As well as this, participants were asked to self evaluate several rating questions regarding the feasibility and ease of use of the strategies, recoded in Table 2.

**Table 1.** Study Results.

| Study Metrics | Participant 1 |  | Participant 2 |  | Participant 3 |  |
| --- | --- | --- | --- | --- | --- | --- |
|  | CCS | AM-MCWN | CCS | AM-MCWN | CCS | AM-MCWN |
| Time to complete maze (minutes:seconds) | **4:00** | 6:43 | 13:28 | **7:26** | 9:41 | **4:00** |
| Number of instructions sent | **72** | 122 | **114** | 116 | 107 | **69** |
| Number of crashes | **3** | 5 | 8 | **5** | 6 | **5** |

**Table 2.** Participant Responses.

| Study Question | Participant 1 |  | Participant 2 |  | Participant 3 |  |
| --- | --- | --- | --- | --- | --- | --- |
|  | CCS | AM-MCWN | CCS | AM-MCWN | CCS | AM-MCWN |
| Difficulty to control the system | 5 | 7 | 5 | 7 | 8 | 4 |
| Accuracy of the system to user inputs | 8 | 7 | 4 | 6 | 3 | 4 |
| Feasibility of implementing this type of control with EPW for people with disabilities | 8 | 8 | 6 | 3 | 3 | 3 |

Ratings were on a scale of 1-10, with 1 being not difficult, not accurate, not feasible at all; and 10 being very difficult, very accurate, and very feasible respectively.

In addition, participants were asked to rate their levels of fatigue, on a scale of 0-10, both before and after the session, and the results of this can be seen in Fig 6 and Fig 7.

**Fig 6.**
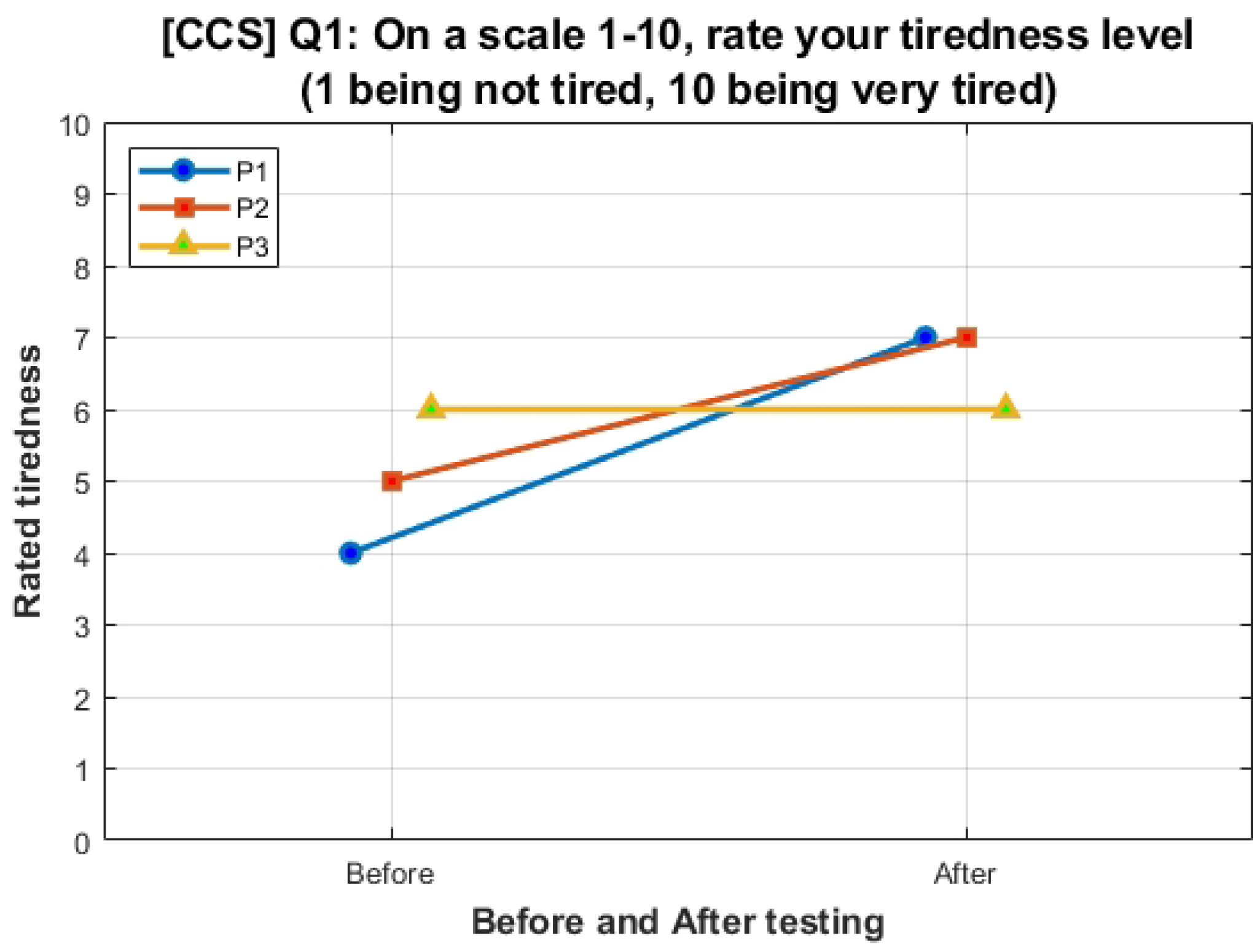
Participant fatigue before and after CCS. Participants were asked to rate their tiredness on a scale of 1-10, 1 being not tired at all, and 10 being very tired.

**Fig 7.**
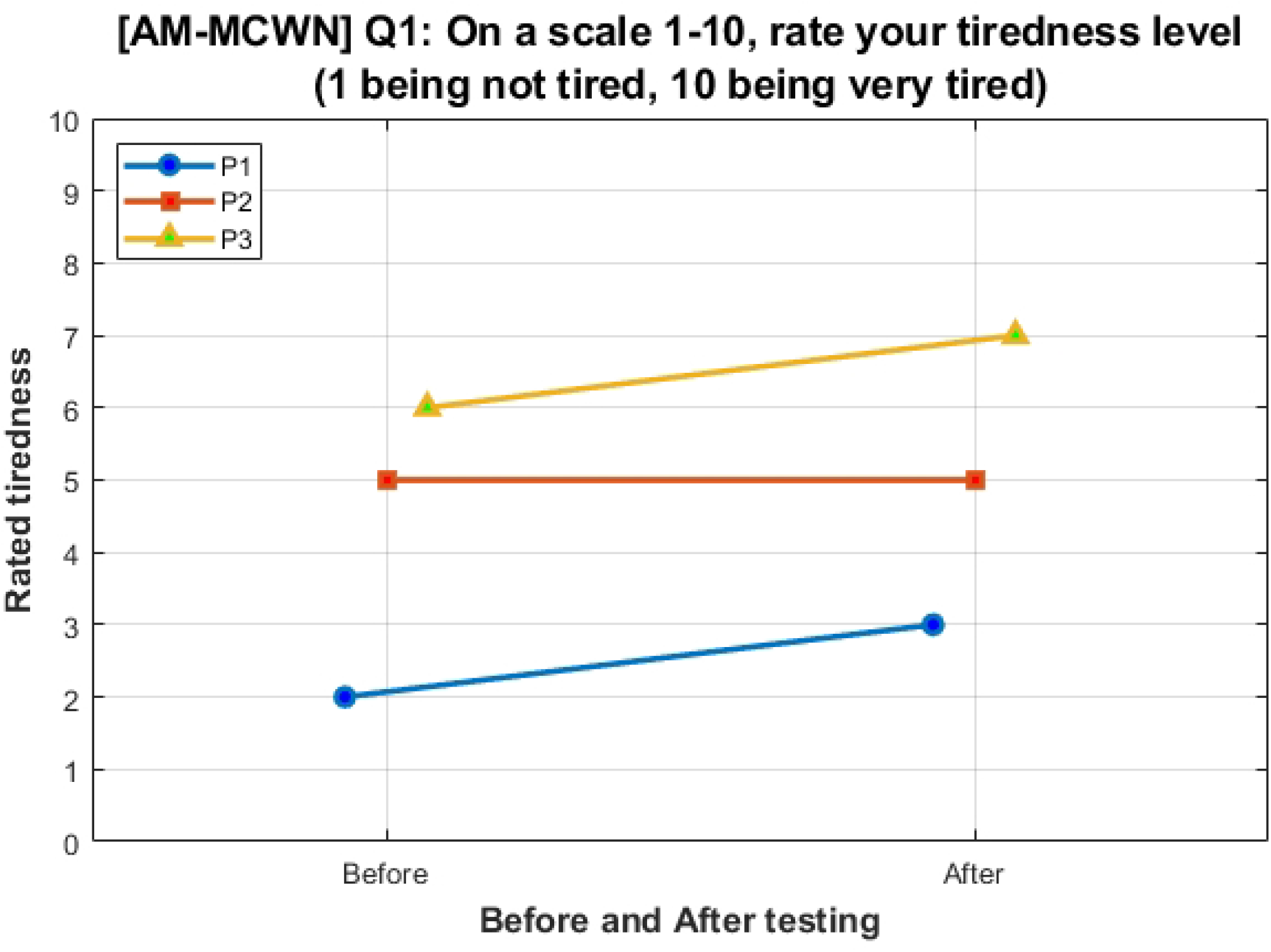
Participant fatigue before and after AM-MCWN. Participants were asked to rate their tiredness on a scale of 1-10, 1 being not tired at all, and 10 being very tired.

It can be seen from the results in Table 1 that CCS had a slightly lower number of operations. Number of crashes and duration to complete the maze was more varied across participants for CCS.

Additionally, participants tended to become more tired after testing CCS, with their tiredness rating sometimes increasing by two points, compared to 0 or 1 for AM-MCWN.

Participants tended to rate AM-MCWN to be more accurate to receiving inputs and more feasible to implement on a real powered wheelchair for disabled users, but more difficult to use.

### Navigation

In addition to metrical values, the physical paths navigated by the participants were recorded using a camera facing the maze and can be seen in Fig 8 and Fig 9.

**Fig 8.**
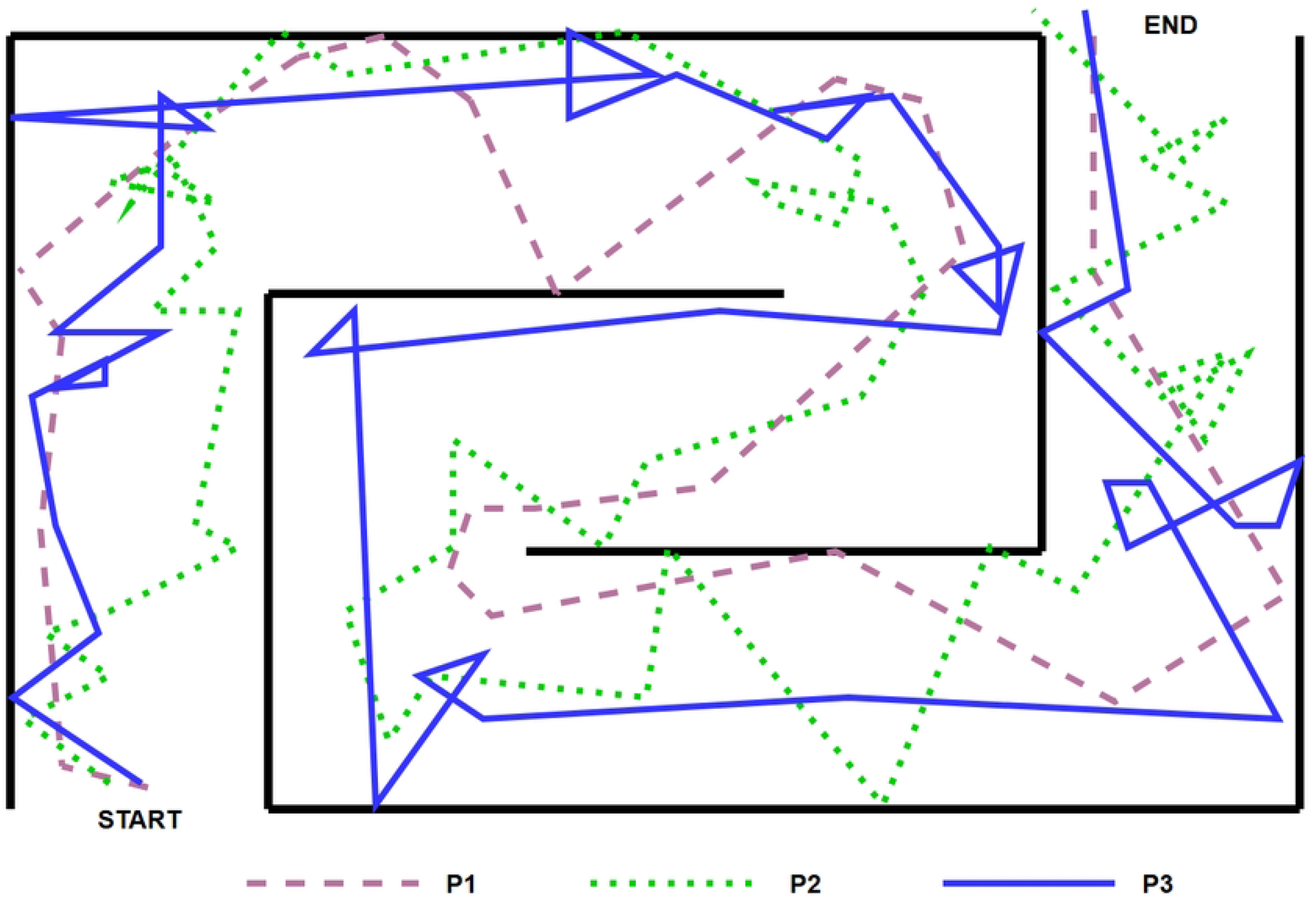
Path travelled by participants during CCS. Recorded path for all three participants navigated by the robotic prototype.

**Fig 9.**
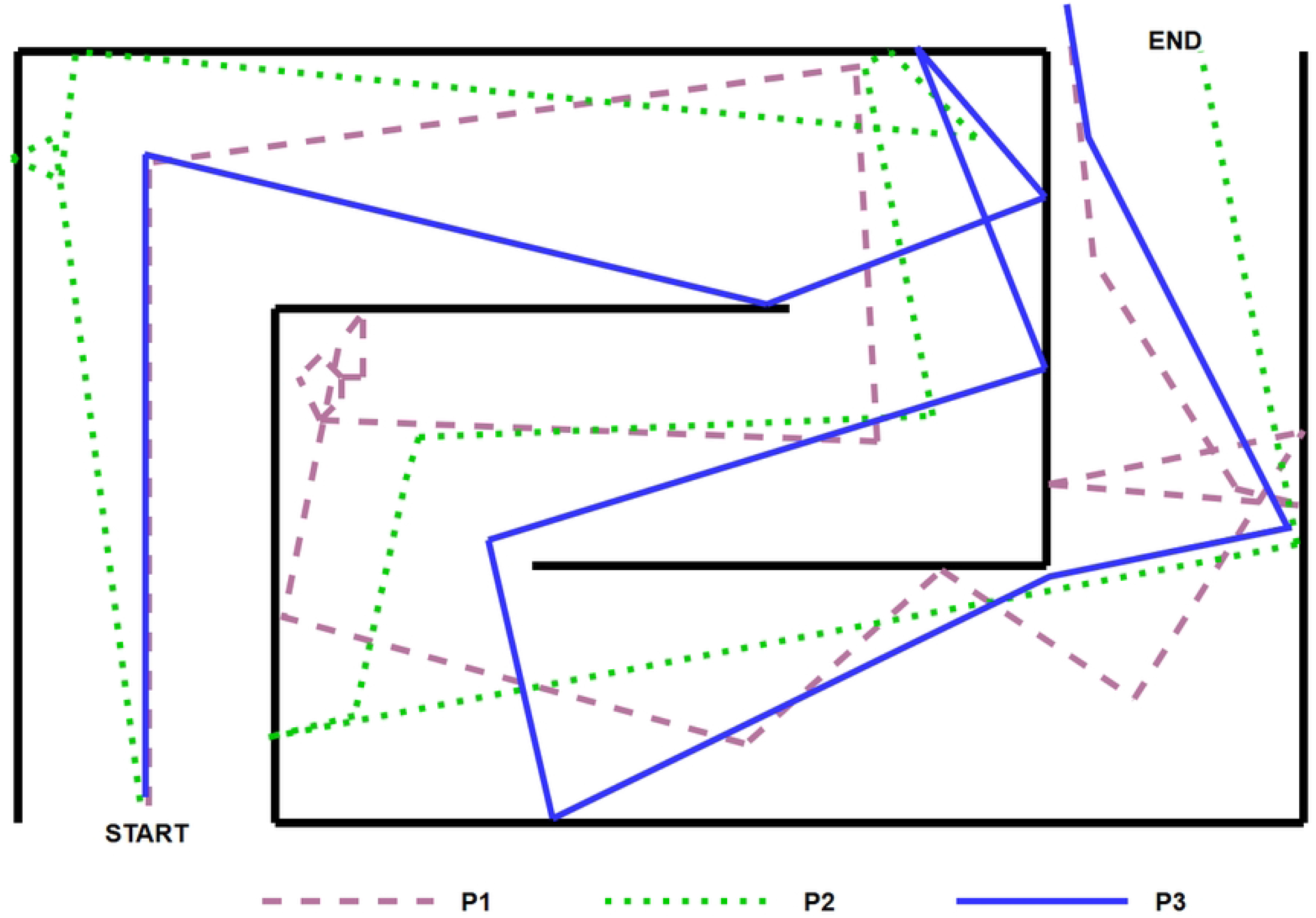
Path travelled by participants during AM-MCWN. Recorded path for all three participants navigated by the robotic prototype.

It should be noted that there were a few points during the continuous run-through, where the wireless sensor lost connection with the ear, and the robot would spin in circles. For both strategies, the user would often switch to reverse mode without meaning to. Both cases would be counted as a single instruction, and the spins may be visualised in Fig 8 and Fig 9 as loops. None of the participants chose to willingly reverse out of crashes, instead opting to turn left and right instead.

### Participant Impressions

Participant 1 mentioned that for AM-MCWN to work well, the user would have to pay attention to the screen: to see what was happening in the system-if a long or short was detected that wasn’t meant to be, and if the user should wait for the system to timeout. They said the same wasn’t true for the CCS, where the user would be making small pulses of the muscle to keep moving in the desired direction, instead of a single long or short signal.

Participant 2 claimed that CCS was more intuitive than AM-MCWN, as the instructions to move left or right were too similar.

Participant 3 claimed that CCS was hard to keep the same (keeping the signal high), though it was more intuitive. They stated that the reverse toggle function was annoying for AM-MCWN, and that it was, in general, too easy to get into for both. They said the delay in CCS was often inconsistent, and sensors often did not pick up the signals very well, though they said the delay could definitely be learnt.

## Discussion

The results from the SVM training demonstrated promising accuracy, with 96% accuracy achieved. Combined with the state machine, timeout function, and immediate stopping, the control strategies are much safer and should be re-tested with a real powered wheelchair to validate their useability.

Overall, both control strategies demonstrated high performance when used to navigate the maze. Both strategies were ranked as average difficulty, which could perhaps be solved with further user training with the AM.

As mentioned in the previous study, and by Schmalfuß et al. [16], perhaps a training program would assist users to become familiar with the use of the AM for control. Users struggled with understanding what can be classified as a ‘long’ or ‘short’ signal, and a training program designed to be a pre-requisite for AM wheelchair control would solve this problem. Additionally, such an investigation would also provide insight as to what user groups can train the AM, and how much. While users can wiggle their ears freely, some can’t move their ears independently left and right, and some cannot keep their facial expressions neutral while doing so. Designing a program that would encourage the users to train their AM would help us understand if all users can be trained to the same level, and if not, then to what level they could be trained to and what systems would be most beneficial for them.

From examining the performance of the two control strategies against eachother, it can be deduced that both strategies may be suitable for different users. For example, AM-MCWN could be used for stroke survivors, as it only requires one EMG set and one muscle group to control, and would be a valuable method for people who only have control of one muscle group they do not regularly use, such as one AM group.

CCS relies on at least two muscle groups working together, with equal amounts of effort from both. This kind of control was rated well and is intuitive to use, so it could be useful for individuals with strong motor skills, as it requires a significant number of inputs. It could be applied to muscles which are more used, such as the forearms.

Additionally, as its default idle state is stop, it makes it the safest system, demonstrated in the results of this study. It would be a practical alternative method of wheelchair control for tetraplegic users, if they were able to train to use their AM independently.

This further stresses the importance of a training program for the AM, as it would be validate that any person would be able to demonstrate learning for the system.

Additionally, the CCS system implemented one electrode on the forearm, and another on the AM, due to participants being unable to move their AM independently. However, if the participants were trained beforehand, they would be able to use the CCS as the author intended.

Participant 3, who had the greatest control over their AM (could move them separately), self evaluated no fatigue increase when using CCS, whereas the other participants reported a more significant increase. This suggests that CCS would be more suitable for more capable AM users.

Alternatively, AM-MCWN did not report as high a change in fatigue before and after using the system, suggesting that the strategy requires less effort to use. However, P1 and P2 reported a higher difficulty of use of AM-MCWN than CCS, perhaps due to its two-command input.

## Limitations

A limitation of the current work is that the control strategy should be tested on a real wheelchair, or at least in a simulation environment, to understand the real-life application challenges with the system, and to remove the additional cognitive workload applied by the user having to perform extra spatial transforms to the prototype. For example, the reverse function may not have been useful in this study setup, where the robotic prototype could turn out of most crashes with a wall, but in a setup with a real user on a wheelchair, participants could be more careful, and end up utilising the reverse function.

In addition to this work, additional research has to be conducted in several other areas, including user investigation, and improvement of electrodes. A major issue during the development and the conduction of the study was the connectivity of the electrodes. The AM area is often an area with a high quantity of hair, therefore the electrodes often loose adhesion and conductivity throughout the study duration. Whenever this would happen, the output of the sensors would short high or low, or oscillate, creating false positives in the classification of the signal-which would cause the prototype to turn, or to spin in circles.

## Conclusion

Many current methods of powered wheelchair control hinder social interaction, thus alternative methods have been explored in recent years. Several studies in current literature have demonstrated and compared implementations for controlling an electric powered wheelchair using EMG control for victims of stroke, amyotrophic lateral sclerosis, and muscular dystrophy.

By comparing two control strategies for the first time, and examining the differences in manoeuvrability, functionality, and their limitations, it can be concluded that some control types will be more suitable to certain applications than others, such as AM-MCWN could be used for victims of stroke, and CCS could be used for tetraplegic users.

The AM-MCWN and CCS have been implemented using the AM to demonstrate improvement and feasibility of using these vestigial muscles for control. The implication of the potential of the use of AM for wheelchair control for users with tetraplegia could be substantial. However, further studies would have to verify the real-time useability of the AM to control a wheelchair, as well as proof that the AM may be trained and as useable for a wide range of users.

As part of this work, a two level SVM classifier was developed to improve the system, by comparing various time analysis windows, before a high classifier accuracy was achieved. The system delay was significantly reduced from 2000ms to approximately 200ms. The most optimal time window was found to be around 300ms, as stated in literature. However, we found that depending on the control strategy on the AM, this may be shorter (200ms for CCS) or longer (300ms, than 500ms for AM-MCWN). This kind of practical comparison and adaptation of window lengths for an EMG-controlled assistive device, has not been done before.

Future work could further investigate the user groups or overcome the difficulties with the current electrode connectivity by developing specialised electrodes for the AM area. Additionally, further work should investigate the training of the AM, to confirm the feasibility of such AM control methods. Alternatively, hybrid methods of wheelchair control could be investigated. Such as, combining MMG or eye-control or blinking with AM EMG. A hybrid system where MMG or ECG is combined with EMG would potentially achieve a higher accuracy than EMG alone, which is supported by literature.

## Data Availability

All study result files and classifier files are available from the Loughborough University Research Repository database (doi: 10.17028/rd.lboro.31399233.)

